# Shedding of infectious SARS-CoV-2 in symptomatic neonates, children and adolescents

**DOI:** 10.1101/2020.04.27.20076778

**Authors:** Arnaud G L’Huillier, G Torriani, F Pigny, L Kaiser, I Eckerle

## Abstract

Children are underrepresented in COVID-19 case numbers, with most pediatric cases exhibiting limited severity, and do not seem to be major drivers of transmission, unlike for other respiratory viruses. That said, SARS-CoV-2 infects children across all age groups, and despite the high proportion of mild or asymptomatic infections, it would be naïve not to consider them as transmitters. To address this point we used cell culture to systematically assess the presence of cultivable SARS-CoV-2 in the upper respiratory tract in a cohort of our institution’s first 23 symptomatic neonates, children and teenagers with COVID-19 diagnosed by RT-PCR.

Median age was 12.0 years (interquartile range [IQR 3.8–14.5], range 7 days-15.9 yrs). Most patients had an upper respiratory tract infection (n=13), followed by fever without source and pneumonia (each, n=2). Samples were collected at a median of 2 days (IQR 1–3) after symptom onset. Median viral load (VL) at time of diagnosis was 3.0×10^6^ copies/ml (mean 4,4×10^8^, IQR 6.9×10^3^–4.4×10^8^) from a nasopharyngeal swab (NPS).

SARS-CoV-2 virus isolation was successful in 12/23 (52%) children after inoculating VeroE6 cells with a NPS specimen. SARS-CoV-2 isolation was determined by the presence of a typical cytopathic effect (CPE) and increased viral RNA in the supernatant. SARS-CoV-2 replication in all positive isolates (12/12) was confirmed by a second passage using new VeroE6 cells.

Virus isolation was successful from NPS from all age groups, with a median initial VL of 1.7×10^8^ copies/ml (mean 7.9×10^8^, IQR 4.7×10^6^–1.0×10^9^) (Figure 1). The youngest patient that SARS-CoV-2 was isolated from was a 7-day old neonate. No correlation between disease presentation and success of virus isolation was observed.

Our data show that initial VLs at diagnosis in symptomatic children is comparable to those in adults, and that symptomatic children of all ages shed infectious virus in early acute illness. Infectious virus isolation success was largely comparable to that of adults, although two specimens yielded an isolate at a lower VL (1.2×10^4^ and 1.4×10^5^ copies/ml) than what was observed in adults.

SARS-CoV-2 shedding patterns of culture competent virus in symptomatic children resemble those observed in adults. Therefore, transmission of SARS-CoV-2 from children is plausible. Considering the relatively low frequency of infected children at this time, biological or other unknown factors could reduce transmission in this population. Both large serological investigations and systematic surveillance of acute respiratory diseases are needed to understand the role of children in this new pandemic.

## Introduction

Children are underrepresented in COVID-19 case numbers ^1,2^, with most pediatric cases exhibiting limited severity, and do not seem to be major drivers of transmission, unlike for other respiratory viruses^3,4^. That said, SARS-CoV-2 infects children across all age groups^1,3^, and despite the high proportion of mild or asymptomatic infections, it would be naïve not to consider them as transmitters. To address this point we used cell culture to systematically assess the presence of cultivable SARS-CoV-2 in the upper respiratory tract in a cohort of our institution’s first 23 symptomatic neonates, children and teenagers with COVID-19 diagnosed by RT-PCR.

## Methods

Our region had a SARS-CoV-2 outbreak, with an estimated 800 cases/100’000 inhabitants^5^. Patients were either seen at the pediatric emergency room of the Geneva University Hospitals, or samples were received by the laboratory from other health care facilities as part of its function as the Swiss national reference laboratory for Emerging Viral Diseases at the Geneva University Hospitals.

All Nasopharyngeal (NPS) specimens were collected with a flocked swab in universal transport medium (both Copan, Italy) and tested for SARS-CoV-2 by according to manufacturers’ instructions on various platforms, including initially in house method using eMAG extraction (bioMérieux, France) and Charité RT-PCR protocol^6^, then BD SARS-CoV-2 reagent kit for BD Max system (Becton, Dickinson and Co, US) and Cobas 6800 SARS CoV2 RT-PCR (Roche, Switzerland). All samples, both extracted RNA as well as remaining original specimens, are stored at −20°C and −80°C, respectively. For this study, RNA extracts of all samples were re-run with the E-gene assay (TibMolBiol, Berlin, Germany) on a Roche Light Cycler 480 (Roche Switzerland) by according to manufacturers’ instructions, by using in vitro transcribed RNA for quantification (European Virus Archive)^7^. Cell culture supernatant was isolated by manual extraction with Machery & Nagel Kit (Düren, Germany) and quantified by the same assay.

For assessment of infectious virus, VeroE6 cells were seeded at a density of 8×10^4^ cells/well in a 24 well plate and inoculated with 200 μl of viral transport medium the following day. Cells were inoculated for 1h at 37°C, then inoculum was removed, cells were washed 1x with PBS and then regular cell growth medium containing was added. Cells were observed on day 2, 4, and 6 for the presence of cytopathic effect by light microscopy. Supernatant was harvested upon the first observation of a CPE, or, if no CPE was observed, at the end of the experiment on day 6. For a second passage, 20 μl supernatant of all CPE positive specimens was transferred onto new VeroE6 cells. Supernatant after inoculation and upon observation of a CPE was collected and isolation of replication competent SARS-CoV-2 confirmed by an increase in viral RNA.

Clinical data of study patients were retrieved after approval by the institutional review board (Commission Cantonale d’Ethique de la Recherche [CCER] protocol 2020–00835) and documented parental consent in the medical charts.

## Results

Among 638 patients <16 years old tested for SARS-CoV-2 by RT-PCR on NPS between January 25 and March 31, 2020, 23 (3.6%) tested positive. Median age of the pediatric patients with positive RT-PCR was 12.0 years (interquartile range [IQR 3.8–14.5], range 7 days-15.9 yrs) (Supplementary Table 1). Most patients had an upper respiratory tract infection (n=13), followed by fever without source and pneumonia (each, n=2). Samples were collected at a median of 2 days (IQR 1–3) after symptom onset. Median viral load (VL) at time of diagnosis was 3.0×10^6^ copies/ml (mean 4,4×10^8^, IQR 6.9×10^3^–4.4×10^8^) from a nasopharyngeal swab (NPS) (Supplementary Table 1).

SARS-CoV-2 virus isolation was successful in 12/23 (52%) children after inoculating VeroE6 cells with a NPS specimen. SARS-CoV-2 isolation was determined by the presence of a typical cytopathic effect (CPE) (Supplementary Figure 1) and increased viral RNA in the supernatant. SARS-CoV-2 replication in all positive isolates (12/12) was confirmed by a second passage using new VeroE6 cells.

Virus isolation was successful from NPS from all age groups, with a median initial VL of 1.7×10^8^ copies/ml (mean 7.9×10^8^, IQR 4.7×10^6^–1.0×10^9^) (Figure 1 and Supplementary Table 1). The youngest patient that SARS-CoV-2 was isolated from was a 7-day old neonate. No correlation between disease presentation and success of virus isolation was observed.

**Figure 1.**
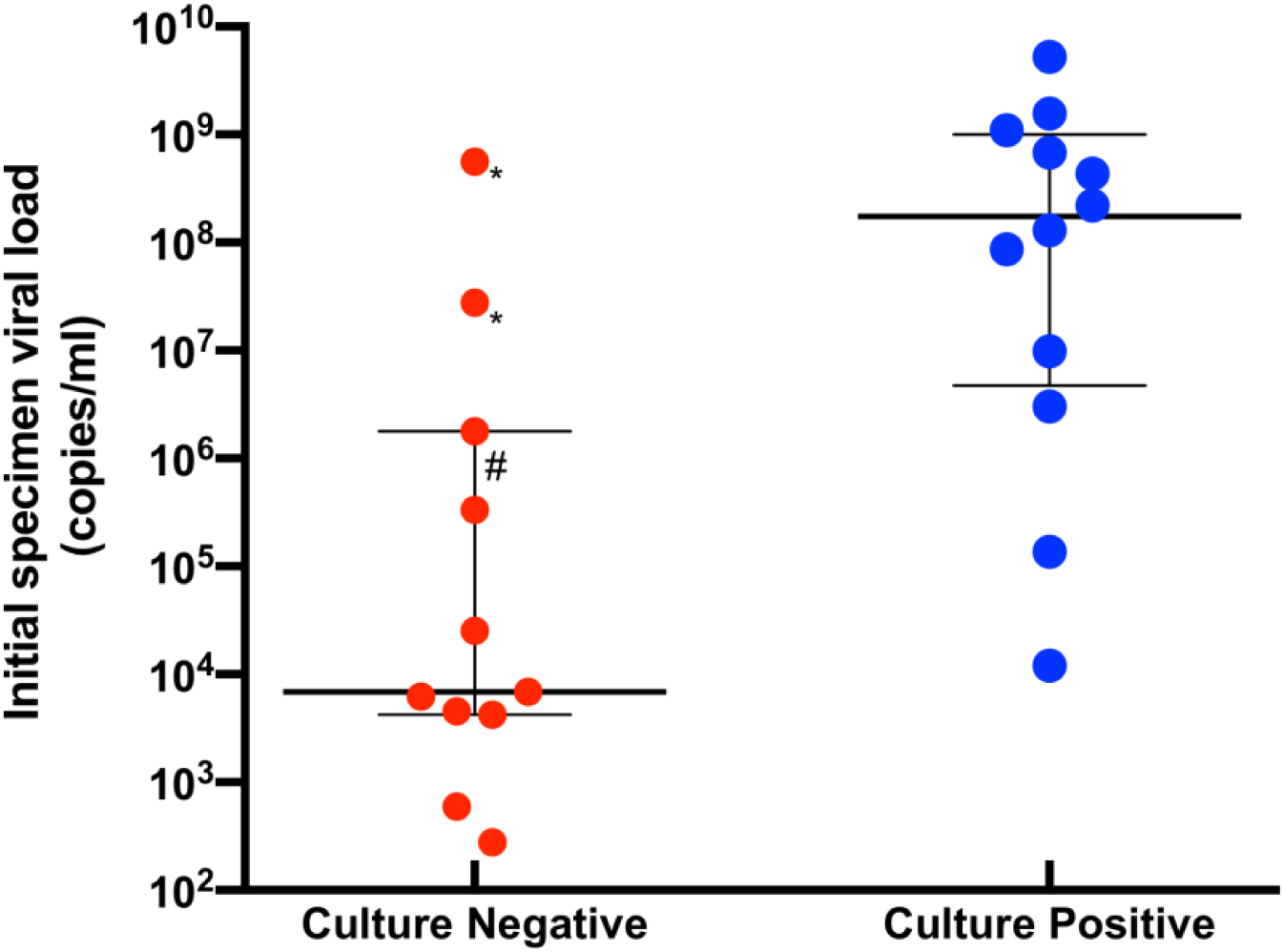
Breakdown of SARS-CoV-2 initial viral load of nasopharyngeal swabs among culture-negative and culture-positive specimens The thick and thin horizontal bars represent the median viral load and the interquartile range, respectively. *: specimen collected outside institution, suggesting a longer time to freezing at −80°C; #: specimen with a duration between collection and freezing at −80°C around 48 hours. SARS-CoV-2: severe acute respiratory syndrome coronavirus 2

## Discussion

Our data show that initial VLs at diagnosis in symptomatic children is comparable to those in adults^8^, and that symptomatic children of all ages shed infectious virus in early acute illness. Infectious virus isolation success was largely comparable to that of adults, although two specimens yielded an isolate at a lower VL (1.2×10^4^ and 1.4×10^5^ copies/ml) than what was observed in adults^8^.

A limitation of our study was the small number of children assessed. Furthermore, the use of left-over material received for routine diagnostic purposes could have resulted in suboptimal times between sample collection and storage at −80°C due to transport and diagnostic processing time, resulting in a loss in infectivity and an underestimation of the initial number of viable viral particles. This might have led to a decreased titer of infectious virus and a failure of virus isolation even in the presence of high viral RNA levels. Of note, 2 of 3 samples with a high viral RNA level but unsuccessful virus isolation were collected outside our institution, and thus had longer transport times to the laboratory; the last one was collected in our institution, but the time between specimen collection and processing was 24 hours. The vast majority of patients were managed as outpatients and had to self-isolate at home after diagnosis, so no consecutive sampling was possible to assess infectious virus in multiple samples over the course of disease.

## Data Availability

Study protocols are available upon request. Proposals should be submitted to

## Supplementary Figure

**Supplementary Figure 1.**
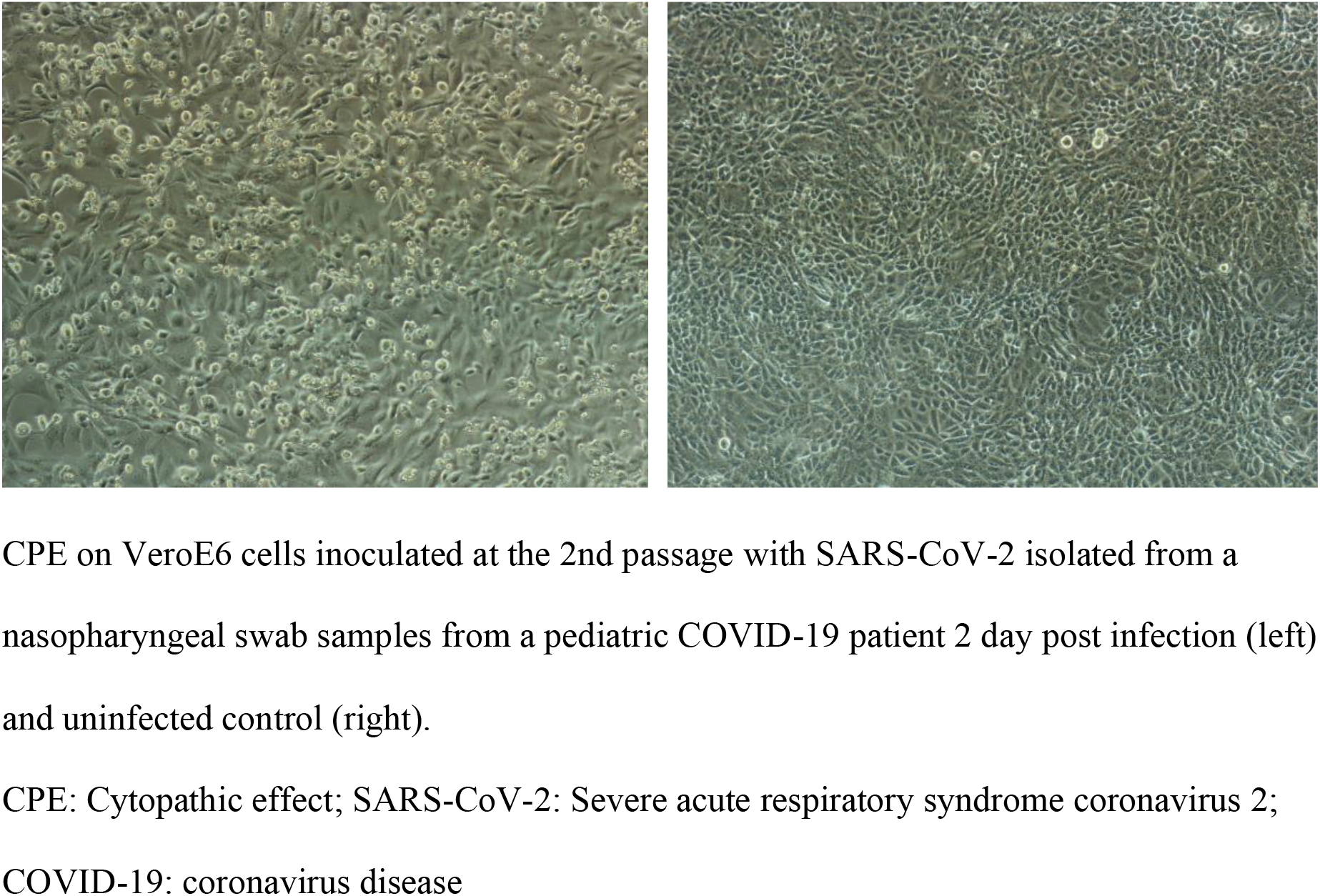
Cytopathic effect on VeroE6 cells inoculated with SARS-CoV-2

**Table S1.** Demographics, initial viral load and culture results of study patients

| **Study Patient** | **Age range** | **Initial specimen viral load (copies/ml)** | **Isolate First passage** | **Isolate Second passage** |
| --- | --- | --- | --- | --- |
| 1 | 10-16 years | 2.8x10<sup>7</sup> | - | no second passage |
| 2 | 5-10 years | 1.8x10<sup>6</sup> | - | no second passage |
| 3 | 10-16 years | 9.9x10<sup>6</sup> | + | + |
| 4 | 10-16 years | 6.9x10<sup>3</sup> | - | no second passage |
| 5 | 1-5 years | 4.5x10<sup>3</sup> | - | no second passage |
| 6 | 10-16 years | 8.6x10<sup>7</sup> | + | + |
| 7 | 5-10 years | 6.2x10<sup>3</sup> | - | no second passage |
| 8 | 10-16 years | 3.3x10<sup>5</sup> | - | no second passage |
| 9 | 3-6 months | 2.8x10<sup>2</sup> | - | no second passage |
| 10 | 1-5 years | 5.9x10<sup>2</sup> | - | no second passage |
| 11 | 5-10 years | 5.6x10<sup>8</sup> | - | no second passage |
| 12 | <1 month | 1.3x10<sup>8</sup> | + | + |
| 13 | 10-16 years | 4.2x10<sup>3</sup> | - | no second passage |
| 14 | 10-16 years | 2.5x10<sup>4</sup> | - | no second passage |
| 15 | 10-16 years | 1.1x10<sup>9</sup> | + | + |
| 16 | 10-16 years | 2.2x10<sup>8</sup> | + | + |
| 17 | 1-3 months | 5.3x10<sup>9</sup> | + | + |
| 18 | 1-3 months | 4.4x10<sup>8</sup> | + | + |
| 19 | 5-10 years | 1.6x10<sup>9</sup> | + | + |
| 20 | 10-16 years | 6.8x10<sup>8</sup> | + | + |
| 21 | 10-16 years | 1.4x10<sup>5</sup> | + | + |
| 22 | 10-16 years | 1.2x10<sup>4</sup> | + | + |
| 23 | 10-16 years | 3.0x10<sup>6</sup> | + | + |

ml: milliliter

## References

1. Livingston E, Bucher K. Coronavirus Disease 2019 (COVID-19) in Italy [published online March 17, 2020]. JAMA 2020. doi: 10.1001/jama.2020.4344.

2. Guan WJ, Ni ZY, Hu Y, et al. Clinical Characteristics of Coronavirus Disease 2019 in China [published online February 28, 2020]. N Engl J Med 2020. doi: 10.1056/NEJMoa2002032

3. Lu X, Zhang L, Du H, et al. SARS-CoV-2 Infection in Children [published online March 18, 2020]. N Engl J Med. doi: 10.1056/NEJMc200507

4. World Health Organization. Report of the WHO-China Joint Mission on Coronavirus Disease 2019 (COVID-19). Published February 28, 2020.

5. Federal Office of Public Health. Accessed April 24, 2020 [Available from: https://www.bag.admin.ch/bag/en/home.html].

6. Corman VM, Landt O, Kaiser M, et al. Detection of 2019 novel coronavirus (2019-nCoV) by real-time RT-PCR. Euro Surveill 2020; 25(3) doi: 10.2807/1560-7917.ES.2020.25.3.2000045

7. European Virus Archive Global (EVAg). Accessed April 24, 2020 [Available from https://www.european-virus-archive.com].

8. Wolfel R, Corman VM, Guggemos W, et al. Virological assessment of hospitalized patients with COVID-2019 [published online April 1, 2020]. Nature 2020. doi: 10.1038/s41586-020-2196-x

